# Development and external validation of the NEO-READY model to predict date of discharge among premature neonatal intensive care patients

**DOI:** 10.64898/2026.01.20.26344137

**Authors:** Hannah Lonsdale, Kevin Patel, Henry Domenico, Ryan S. Moore, Allison B. McCoy, Benjamin French, S. Trent Rosenbloom, Daniel W. Byrne, Robert E. Freundlich, Mhd Wael Alrifai

## Abstract

**OBJECTIVE:** To develop a parsimonious, interpretable, and accurate model for predicting discharge for premature infants in the NICU that is suitable for prospective evaluation and integration into clinical workflows.

**STUDY DESIGN:** Using routinely available electronic health record data, we developed and validated NEOnatal Reliable Estimation of Approaching Discharge in Young infants (NEO-READY), a daily-updating model that predicts likelihood of discharge within 5 days for premature infants.

**RESULTS:** Data from 702 infants were used to develop the model, and data from 201 infants were used for temporal external validation. The model includes 13 predictors and two interaction terms and demonstrated excellent discrimination across development (AUC = 0.88, 95% CI 0.87-0.90) and validation (0.90, 0.88-0.91) cohorts.

**CONCLUSION:** This work represents step 1 toward our long-term goal: integrating the NEO-READY model into clinical workflows as part of a comprehensive strategy to improve discharge preparedness, reduce discharge delays, and optimize NICU resources.

## INTRODUCTION

Premature infants admitted to the neonatal intensive care unit (NICU) often experience prolonged and medically complex hospitalizations due to associated complications such as intraventricular hemorrhage, retinopathy of prematurity, bronchopulmonary dysplasia, and sepsis [1]. These infants account for over half of NICU admissions, and more than 40% require surgical intervention during their stay, further extending hospitalization [1].

Discharge from the NICU is a resource-intensive process requiring early, coordinated planning among physicians, nurses, discharge coordinators, social workers, and case managers, along with completion of caregiver training, follow-up arrangements, and screening examinations [2]. Failure to anticipate discharge readiness can prolong hospitalization unnecessarily, consuming scarce NICU capacity and increasing healthcare costs, while preventing optimal care of critically ill infants [3]. Despite the operational, financial, and clinical importance of timely discharge, the burden and causes of discharge delays in premature infants are not extensively reported. To our knowledge, there has only been one study that examined incidence of delayed discharge from the NICU to home; this study found 116 documented delays accounting for 480 lost bed-days over 2 years in a single unit [4]. Families share this burden: in a survey of late preterm infants’ caregivers, 12% reported being unprepared for discharge, with lower readiness associated with greater difficulty adjusting and higher healthcare utilization, including twice as many primary care calls and urgent care visits [2, 5, 6].

Accurate prediction of clinical readiness for discharge could improve family preparedness and reduce anxiety, facilitate earlier initiation of complex discharge tasks, and increase NICU bed availability. Most published models focus on estimating NICU length of stay or identifying infants at risk of prolonged hospitalization but have limited potential applications to clinical practice and/or discharge preparation. Existing models are often designed only to predict length of stay on the day of admission [7–16], have exclusions that render them unsuitable for prospective use [11, 12, 16–18], or use data that would not be available during the patient’s admission, such as billing codes [19, 20].

Temple and colleagues [17] used progress note data and machine learning to predict discharge within 2-10 days. Their models achieved the highest performance at 2 days before discharge, with an area under the curve (AUC) of 0.854. However, a 2-day prediction window does not allow sufficient time to complete discharge planning for complex patients. Finally, only two published studies report external validation and prospective testing [14, 21], and none report measurable impact on patient care, or progression to clinical implementation.

The objective of this study was to develop and validate a prognostic model that can be applied daily to generate a dynamic score indicating the likelihood of discharge within the next 5 days for premature infants in the NICU. By using readily available data to produce updated daily predictions and by accurately predicting discharge within a 5-day window, the model addresses limitations of prior approaches and aims to facilitate the timely initiation of discharge planning. Our long-term goal is to integrate this model into the NICU electronic health record workflow as part of a collaborative strategy to improve infant, parent, and care team preparedness; reduce discharge delays; and optimize NICU care capacity.

## METHODS

### Study population

This study used retrospective electronic health record (EHR) data from patients born between 28 and 37 weeks of gestation who were admitted for active care to a Vanderbilt University Medical Center (VUMC) NICU after November 2, 2017, and discharged before January 01, 2020. No further inclusion or exclusion criteria were applied. The study period was chosen to begin with the implementation of our current EHR [Epic, Epic Systems Corporation, Verona, Wisconsin, USA] and to end before the start of the COVID-19 pandemic, due to concerns of an exceptional impact on discharge practices and coordination. The development cohort consisted of 702 premature patients who met inclusion criteria and were admitted for active care to the VUMC NICUs between November 2, 2017, and July 1, 2019. The temporal external validation cohort consisted of 201 premature patients admitted for active care to the VUMC NICUs between July 2, 2019, and discharged before January 01, 2020. All hospital identifiers were removed, and patients were assigned a unique study identifier to protect privacy. This study was approved by the Vanderbilt University Medical Center Institutional Review Board [LONSDAH09192024120046].

The VUMC NICUs comprise 119 beds across three geographic areas within the medical center: two private-room units in a free-standing children’s hospital and a third open-bay unit adjacent to the labor and delivery area within the adult hospital. The VUMC NICUs admit approximately 1500 infants annually and serve as a regional perinatal referral center with approximately 40% of patients transferred from other facilities (outborn). The patient population includes preterm and term infants, including those with complex surgical and cardiac anomalies. In keeping with standard practice at most US institutions, infants are routinely discharged directly home from the NICU without transition to stepdown units [22].

### Data collection and processing

We obtained retrospective data from the EHR reporting database (Clarity). Data were processed using Structured Query Language (SQL) to create fields containing one observation per day for each patient. The dataset was then screened for missing and non-physiological values. The latter were removed from the dataset before further processing and treated as missing. Non-physiological values may be introduced as typographical errors when clinical staff manually enter data in a patient’s chart. Wherever possible, missing values were then filled by manual chart review. The degree of true missingness (after chart review) for each variable is reported in Table 1. For vital signs and feeding intake, within-patient last-observation-carried-forward was used to impute missing daily values. Median value imputation was used in cases of first-day missing data for continuous variables, or an “Unknown” category in cases of categorical variables.

**Table 1.** Variables used in the modeling process.

| Input feature | Source | Degree of missingness | Definition |
| --- | --- | --- | --- |
| Gestational age at birth (weeks + days) | Administration | 0 | Time since mother's last menstrual period |
| Birth location (inborn/outborn) | Administration | 0 | Born at VUMC (inborn) or transferred to VUMC after birth (outborn) |
| Current day of admission | Administration | 0 | Number of days since admission |
| Daily weight (g) | Flowsheets | 7.4% | Maximum recorded weight in the past 24 hours |
| Average daily mean arterial pressure (MAP) (mmHg) | Flowsheets | 1.7% | Daily mean of documented MAPs in the past 24 hours |
| Percentage oral intake (%) | Flowsheets | 7.2% | Percentage of documented volume of breastmilk and/or formula administered orally in the past 24 hours |
| Highest environmental temperature (°C) | Flowsheets | 0 | Maximum incubator set temperature, or room temperature (26°C) if nursed in an open crib in the past 24 hours |
| Caffeine administered (yes/no) | Medication administration record | 0 | Yes, if oral or intravenous caffeine was administered in the past 24 hours |
| Significant cardio-pulmonary events (count/day) | Flowsheets | 0 | Count one when any of the following occur in the past 24 hours:<br>• Heart rate <80 for >5 seconds<br>• Oxygen saturation <85 for >5 seconds<br>• Apnea for >20 seconds<br>• Recorded intervention, any of: moderate or vigorous tactile stimulation, PPV, CPAP, blow-by oxygen<br>...and <u>not</u> during feeding, suctioning, or gastric tube placement |
| CRP measured (yes/no) | Laboratory results | 0 | Yes, if any C-reactive protein result in the past 7 days |
| Blood gas measured (yes/no) | Laboratory results | 0 | Yes, if any blood gas result in the past 7 days |
| Medications administered (count) | Medication administration record | 0 | Number of unique medications administered in the past 24 hours |
| Hematocrit measured (yes/no) | Laboratory results | 0 | Yes, if any hematocrit result in the past 7 days |

Features are updated daily at midnight with the previous calendar day's information. Missingness reported at a patient-day level. *VUMC* Vanderbilt University Medical Center, *MAP* mean arterial pressure, *PPV* positive pressure ventilation, *CPAP* continuous positive airway pressure, *CRP* C-reactive protein.

### Outcome

The prognostic model was developed using a discrete-time framework with data at the patient-day granularity. The binary outcome was defined as discharge within 5 days of the prediction date. A 5-day window was considered sufficient time to complete discharge planning for a majority of patients, while allowing enough to be known about the patient’s clinical condition to result in reliable prediction. Patients were censored in the event of death or discharge to a location other than home. To prevent information leakage, once a patient met outcome criteria (discharge within 5 days), subsequent inpatient days were excluded from the analysis.

### Potential predictors

A group of neonatology experts identified relevant features associated with readiness for discharge from published literature and their clinical experience. The list of relevant features was reviewed, pruned, and supplemented in collaboration with informaticians to select only fields reliably available on each patient day. Features were included as candidate predictors if they were available during the hospital admission and could be used to update the prediction daily. Forty candidate predictors (Supplementary Table 1) were assessed. Variables were selected based on their daily availability in the chart, reliability, prior evidence supporting their association with length of stay, clinical plausibility, and their potential to improve model performance. With 638 events in the development cohort, we calculate 16.0 events per candidate degree of freedom, which is sufficient to prevent overfitting.[23]

### Statistical analysis

Variables were summarized as counts and percents for categorical variables and median with interquartile range (IQR) for continuous variables. Univariate associations were tested for using the Mann-Whitney U test for continuous variables and Pearson’s chi-square test for categorical variables. Because data are structured as one observation per patient-day, rather than a continuous time interval, a discrete-time survival model with a binary logistic regression classifier [24] was used. LASSO (least absolute shrinkage and selection operator) with 10-fold cross-validation [25] was used for final variable selection. After variable selection, the final model was fit using a standard generalized linear model. Table 1 lists the final variables that were included in the model.

The prognostic accuracy of the final model was evaluated using measures of discrimination (Area Under Receiver Operating Characteristic Curve (AUC) and Area Under Precision Recall Curve (AUPRC)) and calibration (calibration intercept and slope, visual examination of calibration curve, and Brier score). In accordance with TRIPOD guidelines,[26] we performed an external temporal validation using a non-overlapping cohort admitted in a later time period and evaluated discrimination and calibration without model updating. We acknowledge that geographic validation is a necessary next step and is planned. Model performance was evaluated in both the development and the temporal external validation cohorts. Variable importance was reported using each predictor’s chi-square test statistic from the final fitted model. Within the validation cohort, Youden’s cut point was used to determine the optimal risk cutoff for binary decision rules at 5-day intervals during each patient’s admission. Model sensitivity, specificity, positive predictive value (PPV), and negative predictive value (NPV) were reported using the highest Youden index as a threshold for each time point.

A 2-sided p-value of <0.05 was used to indicate statistical significance. No adjustments were made for multiple comparisons. All analyses were conducted using R version 4.4.0. Reporting followed Transparent reporting of a multivariable prediction model for individual prognosis or diagnosis (TRIPOD) reporting guidelines for prognostic studies [26]. Elements of the PROBAST tool [27] to assess risk of bias and the applicability of prediction model studies were incorporated to facilitate future assessment of fairness and applicability.

## RESULTS

### Patient characteristics

The 702 infants in the development cohort (admitted between November 2, 2017, and July 1, 2019) contributed a total of 17 842 patient-days. The 201 infants in the validation cohort (admitted between July 2, 2019, and discharged before January 1, 2020) contributed 4587 patient-days, and were used to temporally validate the model. The characteristics of the patients in each cohort are summarized in Table 2. A total of 85 (9.4%) patients either died or were back-transferred to other NICUs and were therefore treated as censored. Patients with lengths of stay <5 days (49/903, 5.2%) were included in the cohort. The longest admission was 243 days, and the shortest was 1 day. Race and ethnicity, while not used as inputs to the prognostic model, were missing or unknown for 7.2% and 60.4% of patients, respectively. All other patient characteristic fields did not have missing data.

**Table 2.** Patient characteristics and demographics.

| **Characteristic** | **Sub-categories** | **Overall (N = 903)** | **Development cohort (N = 702)** | **Validation cohort (N = 201)** |
| --- | --- | --- | --- | --- |
| **Length of stay (days)** |  | 19 (12-32) | 19 (11-33) | 19 (13-27) |
| **Gestational age at birth (weeks)** |  | 33 (32-35) | 33 (32-35) | 34 (32-34) |
| **Birthweight (g)** |  | 1 921(1 550-2 340) | 1 920(1 540- 2 380) | 1 950(1 660-2 250) |
| **Sex** | Male | 462 (51.2%) | 362 (51.6%) | 100 (49.8%) |
|  | Female | 441 (48.8%) | 340 (48.4%) | 101 (50.2%) |
| **Transferred in from another hospital (outborn)** |  | 233 (25.8%) | 187 (26.6%) | 46 (22.9%) |
| **Race** | White | 616 (68.2%) | 489 (69.7%) | 127 (63.2%) |
|  | Black | 182 (20.2%) | 135 (19.2%) | 47 (23.4%) |
|  | Asian | 6 (0.7%) | 6 (0.9%) | 0 (0%) |
|  | Other | 34 (3.7%) | 20 (2.8%) | 14 (7.0%) |
|  | Unknown | 65 (7.2%) | 52 (7.4%) | 13 (6.5%) |
| **Ethnicity** | Hispanic | 112 (12.4%) | 84 (12.0%) | 28 (13.9%) |
|  | Not Hispanic | 246 (27.2%) | 190 (27.1%) | 56 (27.9%) |
|  | Unknown | 545 (60.4%) | 428 (61%) | 117 (58.2%) |
| **Preferred language** | English | 812 (89.9%) | 635 (90.5%) | 177 (88.1%) |
|  | Spanish | 64 (7.1%) | 47 (6.7%) | 17 (8.5%) |
|  | Other | 27 (3.0%) | 20 (2.8%) | 7 (3.5%) |
| **Died or transferred to a location other than home (censored)** |  | 85 (9.4%) | 64 (9.1%) | 21 (10.4%) |

Categorical data are shown as the number of patients (percentage). Continuous data shown as median value (interquartile range).

### Model description and performance

The final logistic regression model included 13 variables and two interaction terms (Figure 1, Supplementary Table 2, https://cqs.app.vumc.org/shiny/NICUDischargePrediction). The variables most strongly associated with discharge within 5 days were, in order of importance, percentage of nutritional intake taken orally for the last 24 hours, highest environmental temperature for last 24 hours, day of hospitalization on which the prediction was made, gestational age at birth, and birth location (inborn/outborn). The final model demonstrated excellent discrimination (AUC, 0.88; 95% CI 0.87-0.90). Calibration was strong with some miscalibration at higher quantiles of predicted risk (Brier score = .036, Figure 2A). Day-by-day model performance (Table 3) was exceptional.

**Fig 1.**
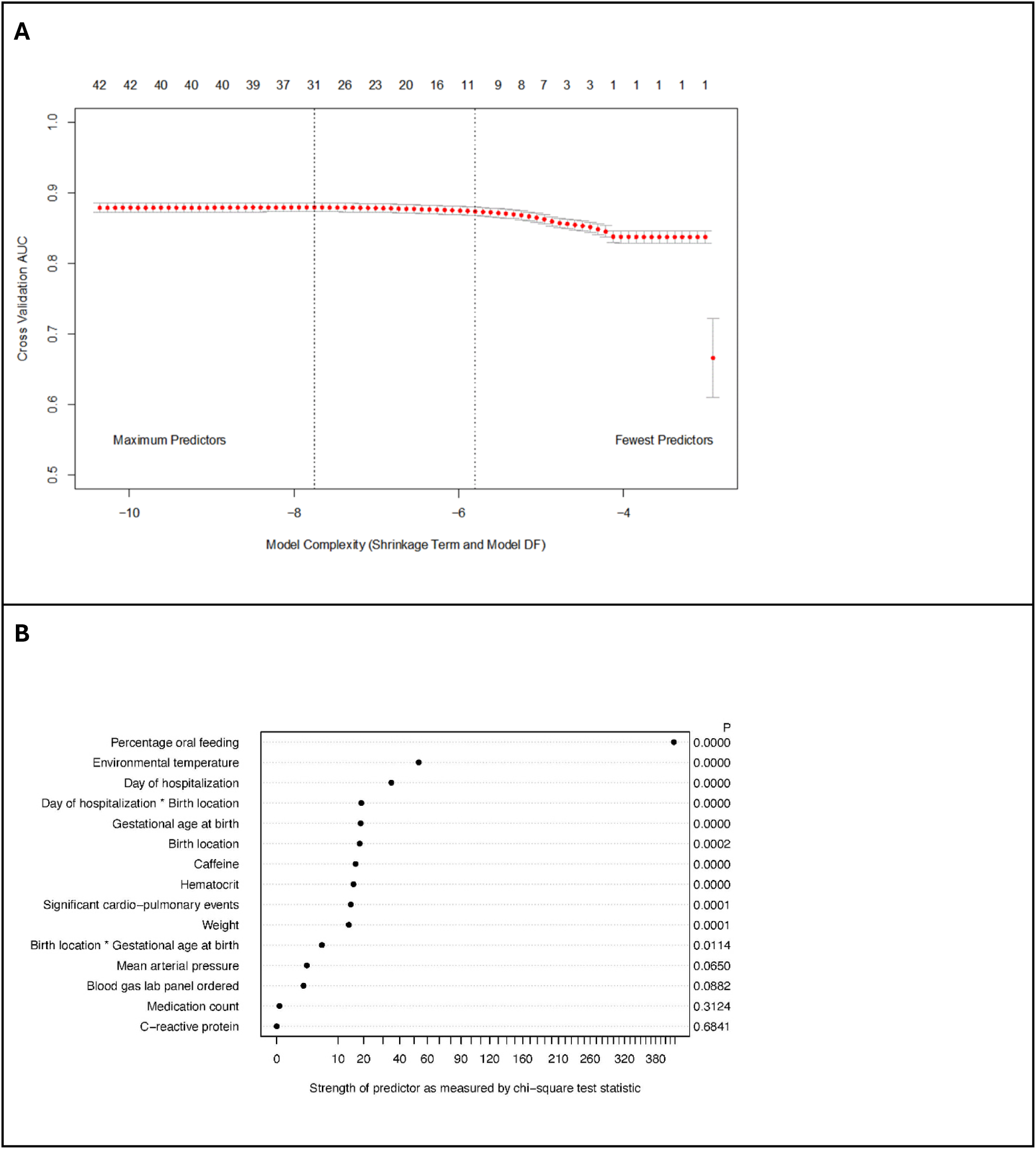
(A) Cross-validation model accuracy vs. model complexity. Accuracy measured as cross-validation AUC with 95% confidence interval. Bottom x-axis shows LASSO shrinkage term. Top x-axis shows model degrees of freedom. **(B)** Predictors included in the final model were ranked by the strength of the predictor, as measured by the model chi-square test statistic. P-values derived from chunk testing of term and subsequent interaction. *AUC* area under receiver operating characteristic curve, *DF* degrees of freedom, *LASSO* least absolute shrinkage and selection operator.

**Fig 2.**
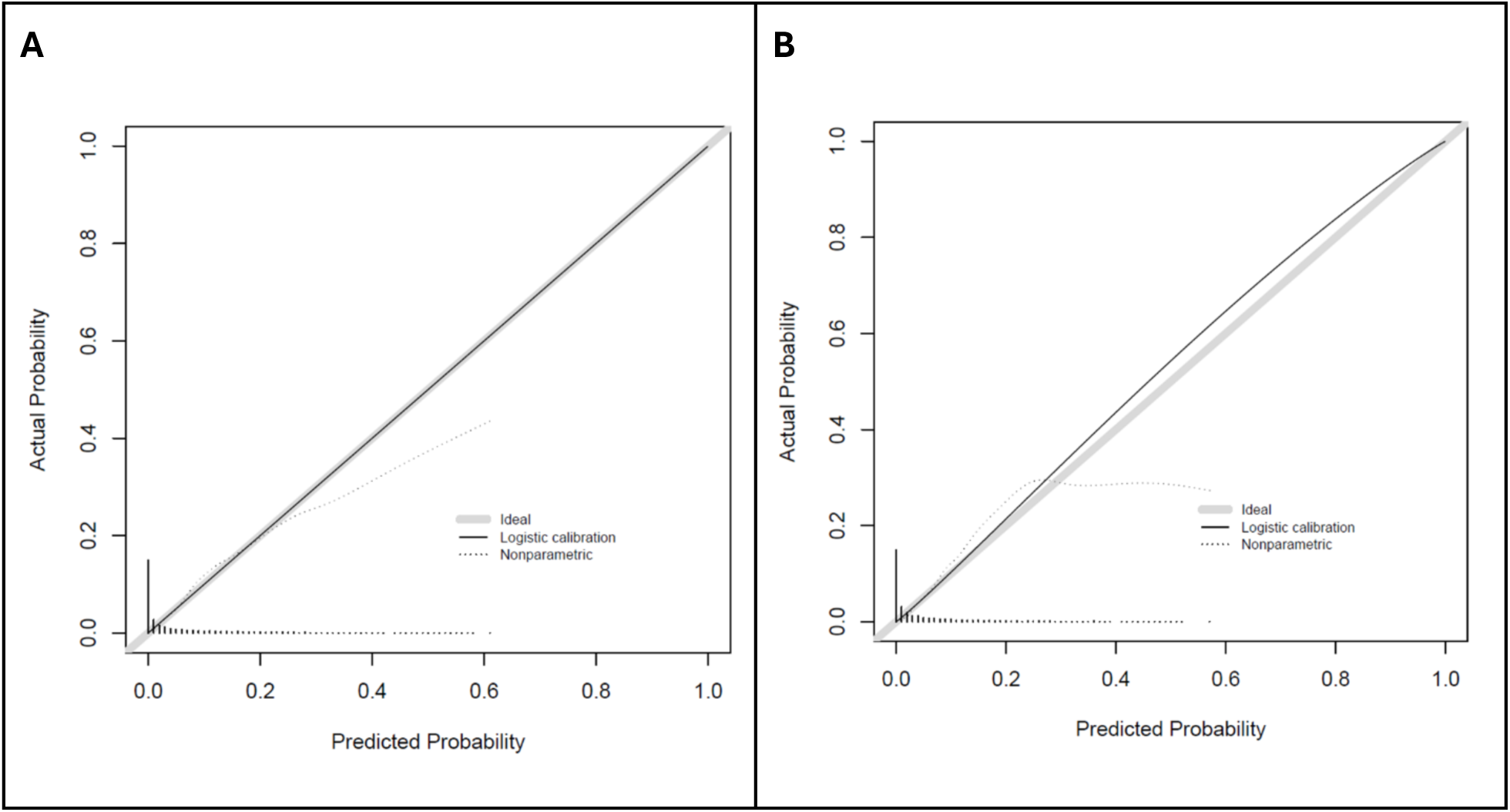
Calibration curves for (A) build cohort and (B) validation cohort.

**Table 3.** Model performance based on the validation set.

| **Day of admission** | **Patients observed (N=201)** | **AUC** | **AUPRC** | **Highest Youden index** | **PPV/NPV** | **Sensitivity/ specificity** |
| --- | --- | --- | --- | --- | --- | --- |
| **5** | **195** | **0.94** | **0.61** | **0.82** | **0.39/1.0** | **1.0/0.82** |
| **10** | **171** | **0.97** | **0.81** | **0.87** | **0.65/0.99** | **0.96/0.90** |
| **15** | **139** | **0.96** | **0.92** | **0.81** | **0.75/0.98** | **0.95/0.85** |
| **20** | **92** | **0.92** | **0.77** | **0.79** | **0.79/0.95** | **0.88/0.91** |
| **25** | **64** | **0.96** | **0.89** | **0.83** | **0.86/0.95** | **0.9/0.93** |
| **30** | **45** | **0.94** | **0.87** | **0.76** | **0.60/1.0** | **1.0/0.76** |
| **35** | **33** | **0.92** | **0.76** | **0.86** | **0.79/1.0** | **1.0/0.86** |
| **40** | **22** | **0.93** | **0.62** | **0.88** | **0.71/1.0** | **1.0/0.88** |

*AUC* area under receiver operating characteristic curve, *AUPRC* area under precision-recall curve, *PPV* positive predictive value, *NPV* negative predictive value.

### Temporal external validation

No significant differences were found in demographic characteristics between the validation and derivation cohorts (Table 2). The model generated from the development cohort was applied to the temporal validation cohort without recalibration, showing excellent discrimination (AUC, 0.90; 95% CI 0.88-0.91) and calibration, again with some expected miscalibration at higher quantiles of predicted risk (Brier score = 0.039, Figure 2B).

## DISCUSSION

We achieved our study’s objective by using temporally separate retrospective datasets to develop and externally validate an accurate prognostic model that can be applied daily to generate a dynamic score indicating the likelihood of discharge within the next 5 days for premature infants in the NICU. The final model, a discrete-time survival model with a binary logistic regression classifier, demonstrated excellent overall discrimination and satisfactory calibration. Input variables were only included if they were readily available and regularly updated in the EHR, to support dynamic, near-real-time predictions. The use of a parsimonious set of 13 input variables makes the model feasible for future automated use at the point of care. Minimal exclusion criteria and censoring patients who were not discharged home, rather than excluding them, also readies the model for future clinical implementation.

The model’s overall performance in the validation cohort is very strong (AUC = 0.90, 95% CI 0.88-0.91), and discrimination improved still further when measured on a day-specific basis (Table 3). This may be because, at the same time during their hospital admission, infants are at a more homogeneous stage of illness, while the overall AUC averages performance over all time points and therefore a more heterogeneous group of clinical conditions. The model generally shows acceptable calibration across the range of clinically relevant risk levels (Figure 2). In the validation cohort, higher predicted probabilities tended to overestimate the observed event rate, which is not unusual given the limited number of high-risk observations. Clinically, management in this group does not depend on small differences within this range, therefore modest over-prediction is unlikely to influence decision-making. We have implemented the model as a Shiny application at https://cqs.app.vumc.org/shiny/NICUDischargePrediction.

Poor anticipation of discharge readiness and failure to complete non-medical preparation before physiological readiness can lead to unnecessarily prolonged hospitalization, which is associated with increased caregiver anxiety [28], excess morbidity and mortality [29], hospital-acquired infections [30], adverse medication events [30], increased cost, and blocking of finite hospital care resources. We intend for our model to support accurate and timely anticipation of physiologic readiness for discharge. Better anticipation of discharge could improve family preparedness, trigger earlier initiation of complex discharge tasks, and enhance NICU resource availability.

Our model is ideally suited for prospective clinical use due to the following characteristics:

- **Feasibility** - Uses readily available EHR data that can be automatically extracted from the EHR without additional user input for daily predictions at the point of care. Only input variables available at the point of daily clinical assessment were included. During daily assessments, we also cannot know which patients will survive or will require back-transfer, therefore those patients were included but censored in the model, rather than excluded from the dataset.
- **Clinically reasonable** - Developed in full collaboration between modeling experts and clinical neonatologists. As a result, all variables have strong clinical validity. A review of the model predictors and their coefficients shows that the first and second most significant predictors, percentage of intake taken orally and highest environmental temperature in the past 24 hours, have the largest contribution to the likelihood of discharge. The ability to take full feeds and to maintain body temperature within external warming devices represent two important criteria for discharge.
- **Applicability** - The inclusion criteria encompass a broad demographic of NICU patients known to be at risk of longer stays and more complex discharge planning needs. However, patients with lengths of stay less than 5 days were intentionally included, despite the potential for negative impact on model performance, as they comprise an essential population of any level III or IV NICU.
- **Dynamic prediction** - Designed to provide continual assessment of each patient’s discharge readiness, updated daily with the patient’s most current clinical information. It is not limited to a specified time or set of input features.
- **Actionable prediction window** - The model produces excellent discrimination for a clinically valuable prediction window. A 5-day prediction window was chosen as an ideal balance between statistical performance and clinical utility, giving sufficient lead-time to complete discharge planning tasks for most patients.
- **Rigorously validated** - A dataset from patients with temporally separate admission dates from the development cohort was used for external validation. The model demonstrated excellent performance on this external validation.

An extensive literature search found several reports of models developed to predict NICU length of stay (Supplementary Table 3). However, only one of these projects [17] was specifically designed for discharge planning. Temple and colleagues [17] used supervised machine learning to predict days to discharge (DTD) for patients in the NICU. Our model demonstrates improved performance compared to that model’s predictions for 4 DTD and 6 DTD (AUCs 0.795 and 0.754, respectively). Additionally, the authors did not report validation beyond the initial training dataset, excluded patient-days with missing data, and used ICD-9 codes assigned after discharge.

Most other published models [7–16] were designed to predict length of stay only at NICU admission. These retrospective, static models are restricted to input data available at admission and therefore cannot account for evolving clinical events that can significantly impact a patient’s length of stay. Such events include sepsis, necrotizing enterocolitis, or failure to wean from respiratory support. Unsurprisingly, all models trained solely using admission data are limited in the granularity of their predictions and therefore have little clinical utility for predicting discharge readiness in individual patients. Additionally, several models are unsuitable for prospective clinical use because they rely on factors such as billing codes, which would not be available at the point of care [19, 20]. Others exclude a large proportion of patients, including those who underwent surgery, were outborn, back-transferred, or had congenital abnormalities [11, 12, 16, 18, 31]. Several studies excluded patients with missing data, rather than attempting chart review or imputation [7, 17, 32], and one used manually collected data [21]. These failings make prospective real-time use impractical or impossible. Finally, our proposed model is one of the few studies found to demonstrate model validation beyond the original dataset [14, 21].

### Limitations

Despite very strong model performance and careful attention to preparation for future clinical use, there are important limitations to consider. The retrospective datasets were collected from a single institution; therefore, the model may not be generalizable to other centers or to present discharge practices at our own institution. Although discharge guidelines are published, each hospital has its own practice and discharge threshold for preterm neonates, which evolves over time and may affect the model’s performance. As with all prognostic models, prospective validation and recalibration as needed for new domains is vital. The model was also trained using actual discharge dates, rather than the date of physiological readiness for discharge. Physiological readiness status is not explicitly recorded at many institutions, including ours. Some patients may have been ready for discharge earlier than their actual discharge date; therefore, the model may perform even better if we can adjust for patients who experience nonmedical delays in discharge.

We trained this initial model to predict discharge for babies born at 28 to 37 weeks of gestation. Term babies were not included, as many are admitted for fewer than 5 days; therefore, the model is of no clinical utility. There is, however, a subset of term infants with complex needs who experience longer stays and may benefit from better discharge forecasting. Babies born at less than 28 completed weeks of gestation frequently have very complex discharge needs and a need for durable medical equipment at discharge; therefore, a 5-day window may not be sufficient to complete discharge preparation. Future work will address extending the prediction window, geographical validation, and exploratory domain validation for extremely preterm and complex term babies.

The model’s discrimination is excellent for lengths of stay of 10 to 40 days (Table 3). Before 10 days, clinical presentations are at maximum heterogeneity. Model performance may be improved by removing patients with a length of stay of less than 5 days, but with prospective use, we could not foresee with certainty which patients would be discharged within 5 days. Complex discharge planning needs are more likely in infants with a length of stay >10 days; therefore, predictions are more useful in this group than for those with shorter admissions. After 40 days, only around 10% of patients remain admitted, limiting the size of the dataset that can be used to train the model. The addition of data from more patients with long stays, or the creation of synthetic training data, may improve model performance on this ‘long tail’.

Development was limited by the availability of structured data within the EHR. Some data points were missing for daily weight, oral intake, and MAP. Although we used established imputation methods, missing data may still introduce bias or inaccuracies in the model. All charted information was accurately transferred to the modeling process, but some EHR data capture is performed by direct observation and so carries the potential for inaccuracy. It can be especially challenging to accurately capture the volume of oral intake during breastfeeding. We therefore used a percentage of total intake, rather than an absolute volume. For clinical implementation, we plan to include imputation methods that can provide the most appropriate real-time replacement for missing values. Completion of race and ethnicity fields is optional in our EHR, and they were not completed for 7.2% and 60.4% of patients, respectively. These fields were not used as input variables, but appraisal of these demographics should be undertaken cautiously for the cohorts.

## CONCLUSION

Despite these potential limitations, the product of our development and validation work is a very strong model to predict discharge from the NICU within 5 days, with clear potential for successful prospective assessment and future implementation. Our model overcomes many limitations of previous work to predict length of stay or discharge readiness for NICU patients. Work is underway to record the date of readiness for discharge for patients at the VUMC NICUs, and this will allow recalibration to predict the date of a patient’s physiological readiness for discharge rather than the actual discharge date. Future work will address prospective temporal validation and exploratory domain validation for complex term and extremely preterm babies, combined with initiatives to embed predictions into the EHR and to limit missing and non-physiological data at the point of capture. We will then demonstrate potential clinical value by directly comparing the model’s predictions to predictions of discharge readiness from attending neonatologists.

Our long-term goal is to integrate this model into NICU workflows as part of a comprehensive strategy to improve infant, parent, and care team discharge preparedness; reduce discharge delays; and optimize NICU care capacity. Successful and sustained improvement in timely patient discharge requires not only excellent forecasting but also action to comprehensively identify and overcome barriers in the current healthcare system. We look forward to further collaborations as we undertake future mixed-methods work leading to a pragmatic randomized controlled trial and clinical implementation.

## ADDITIONAL INFORMATION

**Supplementary information** The online version contains supplementary material available at [PROVIDE URL].

## ETHICS APPROVAL AND CONSENT TO PARTICIPATE

This study was approved by the Vanderbilt University Medical Center Institutional Review Board [LONSDAH09192024120046], who determined that the study posed minimal risk to participants. All aspects of this study were performed in accordance with the Declaration of Helsinki.

## DATA AVAILABILITY

The data underlying this study were derived from electronic health records and contain protected health information. As such, they are not publicly available. Access to de-identified data may be granted upon reasonable request, subject to institutional review and data use agreements. An interactive version of the model can be viewed as a Shiny app at https://cqs.app.vumc.org/shiny/NICUDischargePrediction.

## AUTHOR CONTRIBUTIONS

Dr. Lonsdale conceptualized the project, selected the data for model development, developed the model, reviewed and revised the manuscript, and approved the final version. Dr. Patel conceptualized the project, selected the data for model development, developed the model, reviewed and revised the manuscript, and approved the final version. Mr. Domenico helped conceptualize the project, curated data for model development, developed the model, reviewed and revised the manuscript, and approved the final version. Mr. Moore helped conceptualize the project, curated data for model development, developed the model, reviewed and revised the manuscript, and approved the final version. Dr. McCoy performed and developed the query to obtain the final data set for model development, reviewed and revised the manuscript, and approved the final version. Dr. French helped develop the model, reviewed and revised the manuscript, and approved the final version. Dr. Rosenbloom helped to conceptualize the project, reviewed and revised the manuscript, and approved the final version. Mr. Byrne helped conceptualize the project and curate data for model development, reviewed and revised the manuscript, and approved the final version. Dr. Freundlich helped conceptualize the project, reviewed the final model, reviewed and revised the manuscript, and approved the final version. Dr. Alrifai conceptualized the project, acquired and reviewed the data, reviewed the final model and statistical analysis, reviewed and revised the manuscript, and approved the final version.

## ACKNOWLEDGEMENTS

The authors would like to gratefully acknowledge the advice of Dan France, PhD, MPH, Departments of Anesthesiology, Medicine, and Biomedical Engineering at the Vanderbilt University School of Medicine, for his advice on statistical methodology, and Martha Tanner and Yvonne Poindexter, Department of Anesthesiology, Vanderbilt University Medical Center, for their editorial contributions.

## GENERATIVE AI STATEMENT

The authors used a large language model (ChatGPT) to draft language during the early preparation of this manuscript. After using this tool, the authors reviewed and edited the content as needed and take full responsibility for the content. Every word has been reviewed, and every sentence has been edited for clarity and accuracy. Citations were manually retrieved from traditional sources (e.g., PubMed).

## Supplementary Information

**Supplementary Table 1.** Overview of extracted features.

| Source | Feature |
| --- | --- |
| Demographics | Gestational age at birth, post-menstrual age, sex, ethnicity, race, preferred language |
| Administration | Admission date/time, discharge date/time, length of stay, discharge status |
| Flowsheets | Birth weight, daily weight, length, head circumference, near-infrared spectroscopy (NIRS) readings, oxygen saturation, body temperature, heart rate, mean arterial pressure (MAP), respiratory rate, form of respiratory support, total intake, total output, volume of formula taken orally, volume of breast milk taken orally, formula by feeding tube, breast milk by feeding tube, incubator set temperature, presence of cooling device, set temperature of cooling device, need for tactile stimulation, NPO status |
| Medication administration record | Caffeine, number of medications administered |
| Laboratory and imaging results | White cell count, red cell count, hemoglobin, hematocrit, platelets, blood culture, sodium, potassium, bicarbonate, blood urea nitrate, creatinine, calcium, glucose, troponin, echocardiogram |
| Clinical notes | Social work, cardiology, neurosurgery, operative report |

*NPO* nil per os.

**Supplementary Table 2.** Full model and coefficients.

| Model Variable | Coefficient | P-value |
| --- | --- | --- |
| Percent of nutritional intake taken orally | 3.2327 | <0.0001 |
| Environmental temperature | -0.1498 | <0.0001 |
| Gestational age at birth | 0.1675 | <0.0001 |
| Birth location | 5.3529 | 0.004 |
| Caffeine administered | -1.2807 | <0.0001 |
| Hematocrit measured | -0.4354 | <0.0001 |
| Significant CPE | -0.0342 | <0.0001 |
| Daily weight | 0.0004 | 0.0001 |
| Birth location: outborn and Gestational age at birth interaction | -0.1332 | 0.0114 |
| Mean daily MAP | 0.0112 | 0.065 |
| Blood gas measured | -0.2702 | 0.088 |
| Medication count | -0.0261 | 0.312 |
| CRP measured | 0.0572 | 0.684 |

The model additionally includes terms for non-linear day of hospitalization and interaction between non-linear day of hospitalization and birth location.
Coefficients for non-linear terms are not directly interpretable, so these terms are not included in the table.
*CPE* cardiopulmonary events, *MAP* mean arterial pressure; *CRP* C-reactive protein

**Supplementary Table 3.**
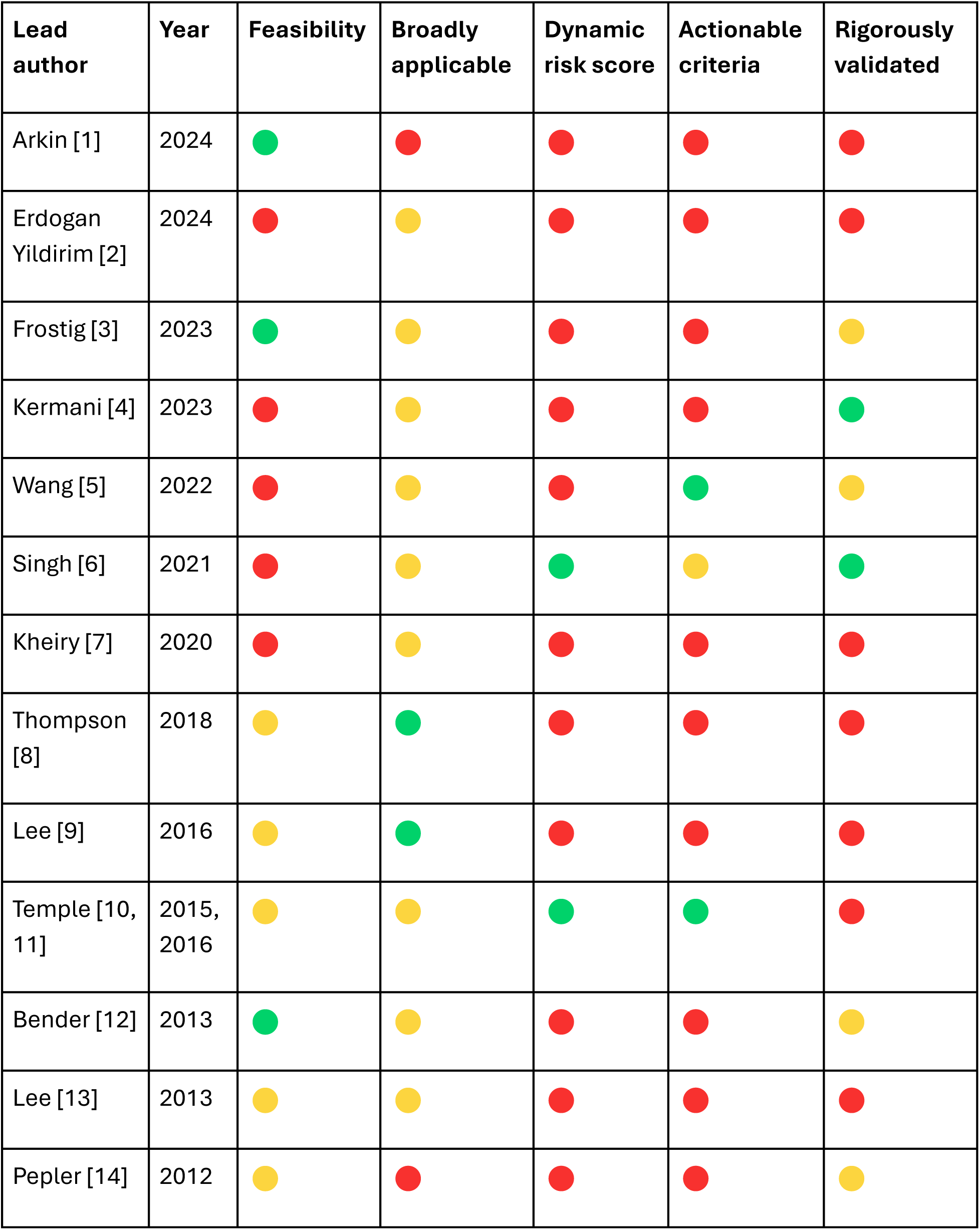
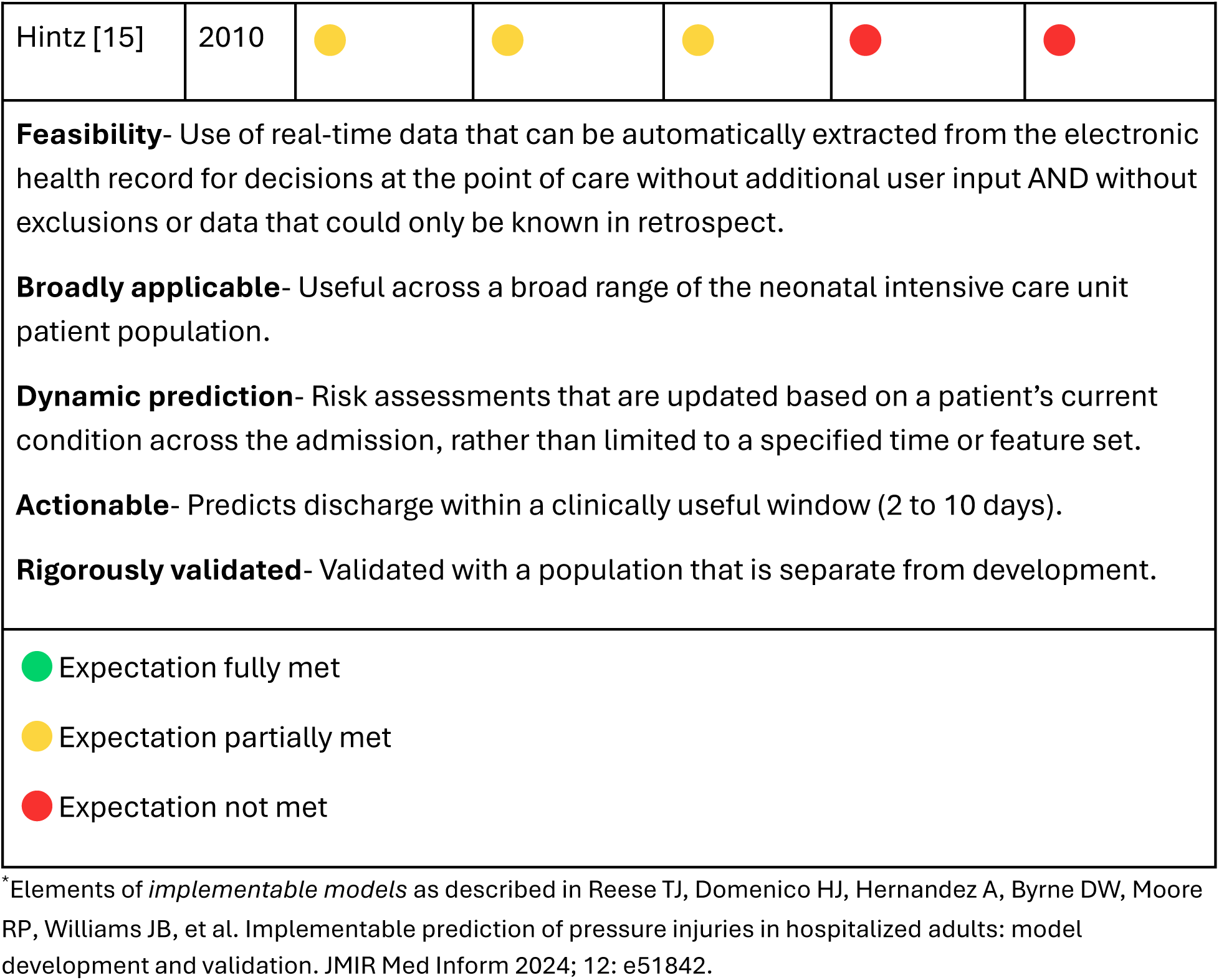
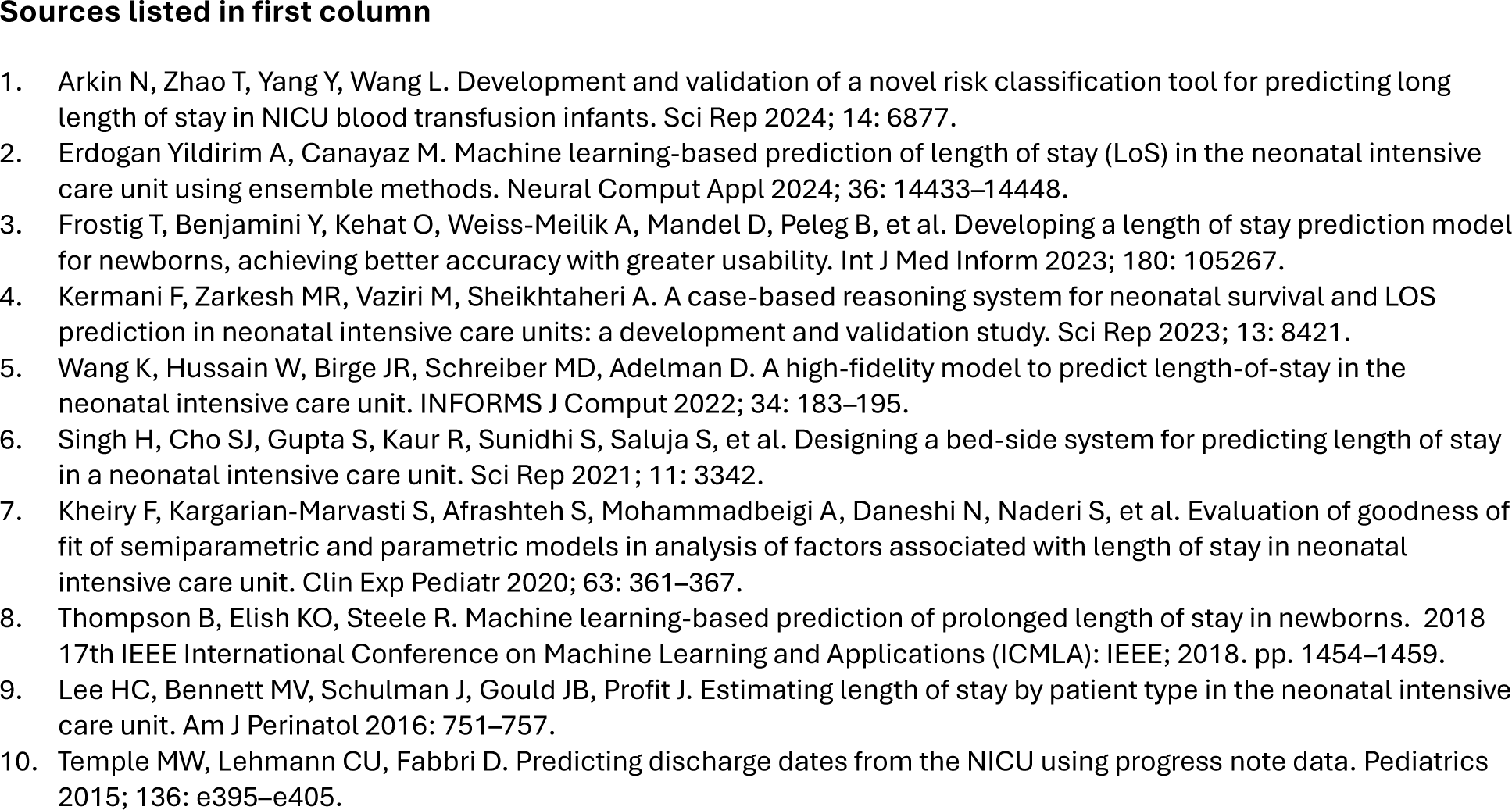
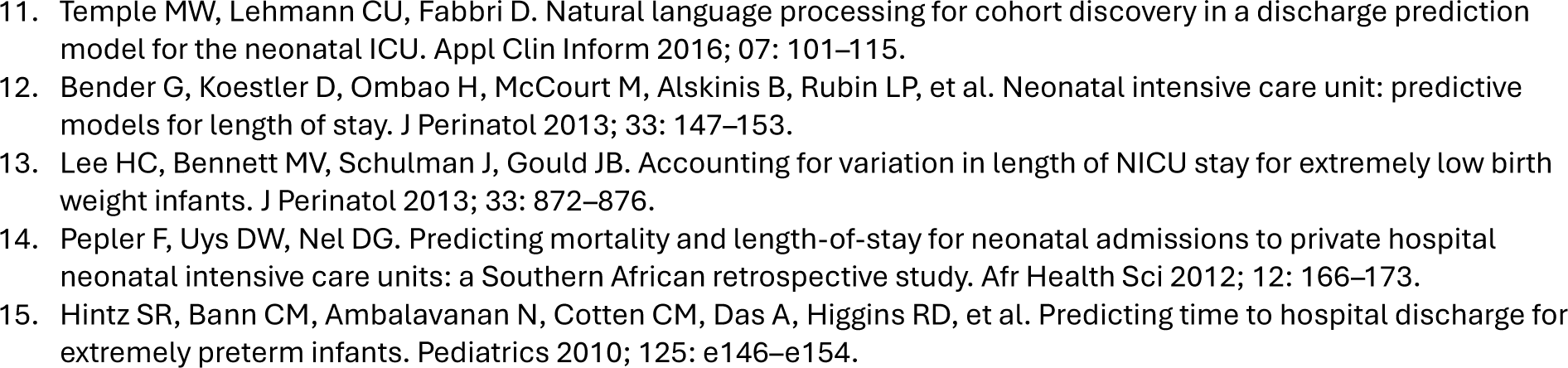
Comparison of current neonatal intensive care unit length of stay and discharge prediction models according to elements of implementable models.* (Arranged in reverse chronological order and limited to publications since 2010.).

